# The Effects of 6 Weeks of Resistance Training on the Gut Microbiome and Cardiometabolic Health in Young Adults with Overweight and Obesity

**DOI:** 10.1101/2023.01.27.23285016

**Authors:** John M.A. Cullen, Shahim Shahzad, Jill A. Kanaley, Aaron C. Ericsson, Jaapna Dhillon

**Author notes:** CORRESPONDING AUTHOR Jaapna Dhillon, PhD Division of Food, Nutrition, and Exercise Sciences College of Agriculture, Food, and Natural Resources School of Medicine University of Missouri, Columbia 312 Gwynn Hall Columbia, MO 65211.

## Abstract

**INTRODUCTION:** Obesity is a known risk factor for the development of insulin resistance and other cardiometabolic disorders. Recently, the gut microbiome has been associated with obesity and subsequent health complications. Exercise has been regularly utilized as a therapeutic intervention to treat obesity and its associated comorbidities. This study examined the effects of a 6-week resistance training exercise program (RT) on the diversity, composition, and metabolic pathways of the gut microbiome.

**METHODS:** Sedentary young adults (age 18-35 years) with overweight and obesity (BMI 25-45 kg/m^2^) were recruited to participate in this randomized controlled trial. Participants were randomized to RT (n=16), a 6-week resistance training program (3 days/week), or control (CT) (n=16), a non-exercising control. Main outcomes of the study included gut microbiome measures (taxa abundances, diversity, and predicted function) and cardiometabolic outcomes (blood pressure (BP) and glucoregulation).

**RESULTS:** Increased abundances of *Roseburia*, a short chain fatty acid (SCFA) producer and predicted starch and sucrose metabolism pathway were observed over 6 weeks (W6) with RT in comparison to CT (group × week, p<0.05, q<0.25). RT also induced higher abundance of microbial flagellar assembly, a pathway involved in cell motility, at W6 compared to CT (group × week, p<0.05, q<0.25). Moreover, RT resulted in higher QUICKI and lower diastolic BP at W6 compared to CT (BL-adjusted p<0.05).

**CONCLUSION:** This study provides preliminary evidence that resistance training increases abundance of selected SCFA producers and microbial pathways and improves cardiometabolic health. Additionally, the associations of cardiometabolic indicators with the microbial community structure warrants further investigation.

## I. INTRODUCTION

The gut microbiome is associated with obesity and related cardiometabolic health disorders (1, 2). This link is primarily supported by animal studies that demonstrate an increased capacity for energy harvesting in an obese gut microbiome as well as a distinct state of dysbiosis characterized by a high *Firmicutes/Bacteroidetes* ratio (1). Additionally some human studies have linked impaired microbial states to metabolic impairments and obesity (3). Direct gut microbial changes are observed with bariatric surgery-induced weight loss in humans with a decrease in weight being positively associated with an increase in microbial diversity following surgery (4). Taken together, these data suggest that the gut microbiome plays a role in obesity etiology and may represent as a therapeutic target for obesity treatment.

Exercise training has been regularly utilized as a therapeutic intervention to treat obesity and its associated comorbidities (5) and recently training has been shown to influence the gut microbiome (6). Alteration of the gut microbiome through exercise training is seen in populations with obesity and lean populations, which appear to reverse after cessation of exercise (7). These changes in the microbiome, while typically observed in long-term exercise interventions occurring over the span of multiple weeks, have also been observed acutely after single bouts of exercise in marathon and ultra marathon runners (8–10), indicating the quick responsive capabilities of the gut microbiome and its response to physical stress (11). Most studies have observed the effects of aerobic exercise (12–19), or combined aerobic and resistance training (15, 20–25) on the gut microbiome while fewer studies examined the effects of only resistance training (RT) (26–29). Unfortunately, numerous lifestyle factors (i.e., diet) and large individual variability observed in the human microbiome have led to conflicting findings. Further, lack of control groups in previous work makes it difficult to attribute any changes to exercise training or other lifestyle changes (14–16, 21, 26–28, 30).

Among the few studies that have utilized RT, some included additional dietary interventions. For example, RT with protein supplementation increased *Veillonellaceae, Akkermansia,* and *Eggerthellaceae*, however the confounding effects of increased protein intake, do not allow, the microbiome changes to be attributed solely to RT (26). Other studies have compared RT to other exercise modalities. In one study, brisk walking was associated with increased intestinal *Bacteroides* while trunk muscle training exhibited no effects on the microbiome in elderly women (30). No control group or full body exercise programming was included in this study. In another study, changes in microbial alpha-diversity were observed with aerobic exercise but not with RT (31). However, in a rat study, RT increased alpha-diversity, SCFA producer *Coprococcus*, and decreased *Pseudomonas, Serratia,* and *Comamonas* genera (29).

Due to the paucity of well controlled trials, the research concerning the impact of resistance training on the gut microbiome in humans is limited. The current study sought to examine 1) the effects of RT on gut microbial diversity, taxa abundances, and predicted function in young adults with overweight and obesity, and 2) to examine the association of the microbiome with cardiometabolic outcomes in this context.

## II. METHODS

This study was approved by the Institutional Review Board of the University of Missouri – Columbia and the trial was registered on clinicaltrials.gov (**NCT04906525**).

### Participants

Young male and female sedentary adults (n=33) with overweight and obesity were recruited for this study and 27 participants completed the study. The participant flow is depicted in **Figure 1**. Twenty-seven participants completed the study. Inclusion criteria included the following: (a) 18–35 years of age, (b) BMI: 25-45 kg/m^2^, (c) sedentary lifestyle, (d) weight stability (≤ 4 kg weight change over the past 3 months), (e) consistent dietary patterns, (f) non-smoker over the previous year, and (g) physically capable of participating in a RT program.

**Figure 1.**
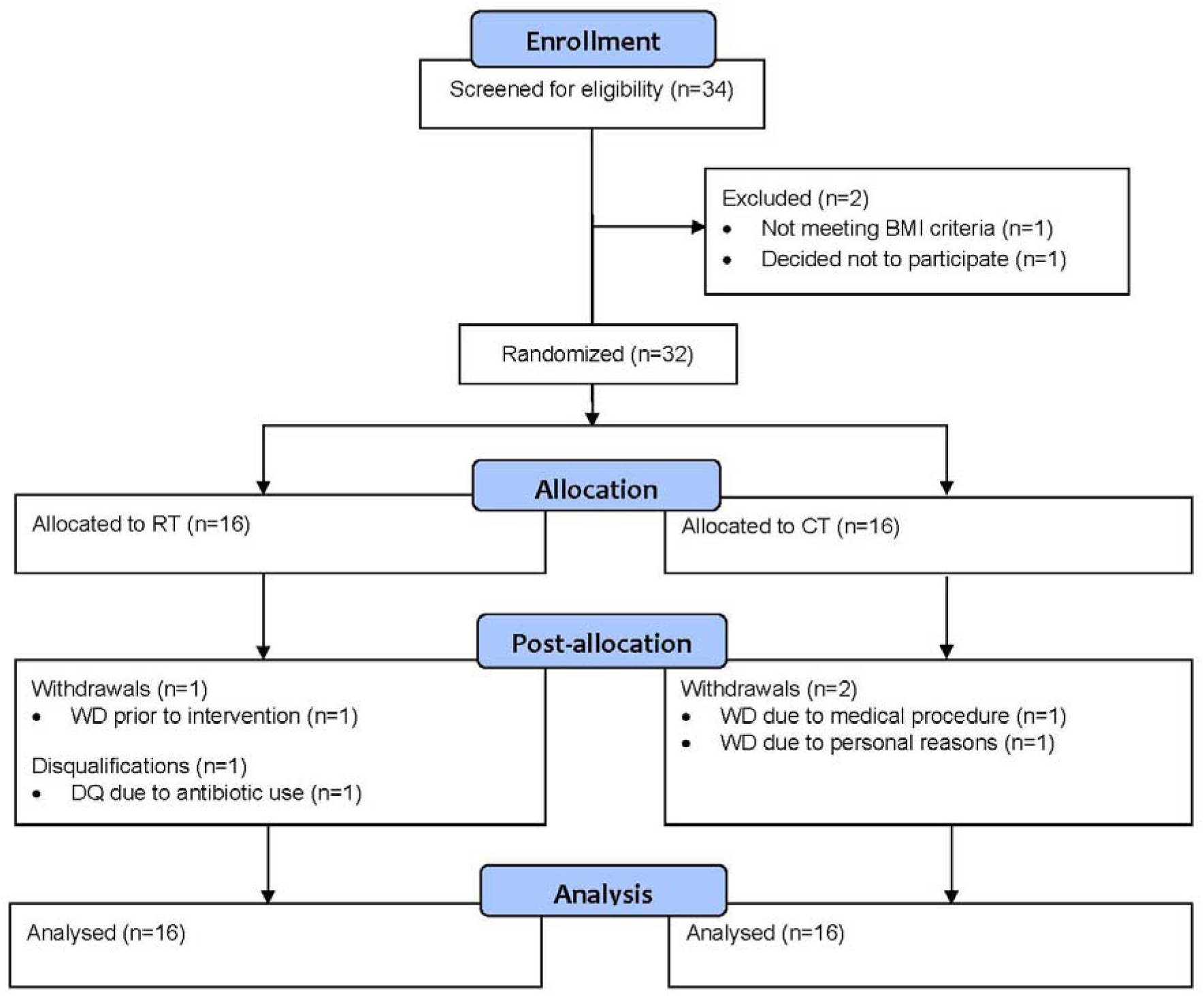
Consort flow diagram of RT and CT from screening to study completion.

Participants with diabetes, uncontrolled hypertension, gastrointestinal diseases or bariatric surgery, pregnancy, use of antibiotics in the past six months, or drug therapy for coronary and/or peripheral artery disease, congestive heart failure or dyslipidemia were excluded.

A health history questionnaire was used to assess medical history and screen for diseases or medications. The International Physical Activity Questionnaire (IPAQ) was used to assess sedentary behavior (32). Those who reported spending most of their days sitting were classified as sedentary. The Physical Activity Readiness Questionnaire (PAR-Q) was used to ensure participants could safely participate in an exercise program by asking questions about past medical history, family medical history, injuries, and symptoms that may be representative of cardiac issues (33). If participants met inclusion criteria, they were invited to an in-person screening visit.

Sample size calculation was done based on based on alpha-diversity outcome from previous dietary intervention studies (34). A power analysis revealed that a sample size of 15 subjects per group is needed to detect 15% change in alpha-diversity with an alpha = 0.05 and power = 0.80.

### Study Protocol

The study was a 6-week randomized, controlled, parallel-arm intervention. Participants were randomized upon arrival to the baseline visit (BL) into 1 of 2 groups: the control group (CT, n=16) or the resistance training group (RT, n=16) using simple randomization via a random number generator in excel. Participants baseline demographic and clinical characteristics are described in **Table 1**. After BL data collection, participants participated in their respective interventions described below and returned at Week 6 (W6) for final data collection.

**Table 1.** Baseline demographic characteristics of participants in the RT and CT groups.

| Characteristics* | RT (*n* = 16) | CT (*n* = 16) |
| --- | --- | --- |
| Sex, *n* (%) |  |  |
| Male | 6 (37.5) | 4 (25.0) |
| Female | 10 (62.5) | 12 (75.0) |
| Age, years | 28.4 ± 4.1 | 28.5 ± 4.4 |
| Race/Ethnicity, *n* (%) |  |  |
| Hispanic | 1 (6.3) | 0 (0.0) |
| Asian/Pacific Islander | 2 (12.5) | 3 (18.7) |
| African American | 2 (12.5) | 1 (6.3) |
| Caucasian White | 11 (68.7) | 12 (75.0) |
| BMI, kg/m<sup>2</sup> | 31.17 ± 4.79 | 31.26 ± 4.32 |
| BMI Category, *n* (%) |  |  |
| Overweight | 8 (50.0) | 6 (37.5) |
| Obese | 8 (50.0) | 10 (62.5) |

\*Data only presented for participants that completed the baseline visit

#### 1. Resistance training intervention

The RT group underwent a 6-week RT intervention consisting of 18 sessions (3 days per week) plus a familiarization session and a final assessment session after W6 data collection (20 total sessions). Participants were instructed to make no changes to dietary habits over the course of the 6 weeks. After completing the BL visit, participants attended the familiarization/assessment session to assess mobility, core strength, and muscular endurance.

Additionally, participants were taught how to properly perform the following exercises: bench press, squat, and barbell row. These three exercises, in addition to plank hold and max push up test, were assessed over the course of the 6 weeks. Participants performed two 3 rep-max (RM) tests for bench press, squat, and deadlift; the first during the first week of programming and the second, after completion of the 6-week intervention. Starting workload was 50-75% of 1RM and was determined by American College of Sports Medicine and National Academy of Sports Medicine trained facilitators. Progression in workload occurred when the participant was successfully able to perform 3 sets of 12 repetitions with proper form. Once workload was increased, repetitions were decreased to 8 until the facilitators determined the participant was capable of increasing repetitions. The facilitator made constant observations to ensure the participant used proper form and was being properly challenged by the given weight. Any necessary modifications to lifts were made by facilitator to ensure the participant could properly perform the lift without risk of injury.

#### 2. Control intervention

Subjects randomized to CT were provided a handout describing the recommended levels of physical activity for their age group but were not given any further instruction or guidance regarding physical activity. They were also instructed to maintain their normal dietary habits and not make any major lifestyle alterations that may influence measured outcomes.

### Study Outcomes and Assessments

#### 1. Microbiome outcomes

Participants were provided stool collection kits at BL and W6 and were also instructed to record their food intake 3 days prior to the BL sample collection. The stool collection kits included sterile collection materials as well as a cooler and ice packs to keep the sample cold until it could be returned to the researchers. Once collected, participants were instructed to keep the sample on ice and return the sample within 24 hours of collection. Once returned to the researchers, the stool sample was aliquoted and frozen at -80 degrees until analysis. Participants were asked to replicate the 3-day diet from their BL sample prior to the W6 sample collection to control for the influence of diet on the gut microbiome. The BL sample was collected prior to the BL study visit. The W6 stool sample was collected and brought to researchers within 48 hours of the final training session after replication of the 3-day dietary log completed at BL.

##### 1.1. DNA extraction and sequencing

DNA from the fecal samples was extracted using Qiagen PowerFecal Kits by the University of Missouri Metagenomics Center as previously described (35). The library preparation and sequencing of the extracted fecal DNA was conducted at the University of Missouri Genomics Technology Core using methodology described previously (36, 37). The V4 region of the 16S rRNA gene was amplified with universal primers (U515F/806R). Paired end reads (2×250 bp) were generated on the MiSeq instrument using Illumina’s standard sequencing protocol.

##### 1.2. Sequency quality control

The demultiplexed paired-end sequence reads from the Illumina platform were imported into QIIME 2 (version 2021.08), a microbiome analysis platform for further analysis (38). The forward and reverse sequences were trimmed i.e., adaptors removed using the cutadapt plugin in QIIME2 (39). The paired-end sequences were filtered, denoised, dereplicated, and merged with the DADA2 quality control package in QIIME2 (40). The amplicon sequence variants (ASVs) obtained from DADA2 were used in all downstream analyses. A total of 4192 ASVs were detected.

##### 1.3. Taxonomy

The ASVs were assigned taxonomy in QIIME2 using a naive Bayes classifier, which was trained on the V4 (515–806) region of the Greengenes 13_8 database reference sequences clustered at 99% sequence similarity (41).

##### 1.4. Microbial pathways

The functions of the microbial community were predicted from 16S rRNA data using Phylogenetic Investigation of Communities by Reconstruction of Unobserved States (PICRUSt2) which infers pathway abundances of gene families such as KEGG orthologs (KO) (42).

##### 1.5. Alpha- and beta-diversity

Alpha-diversity measures assessed included: 1) Chao1 index to assess microbial richness (43), 2) Shannon index to assess both evenness and richness (44), 3) Simpson’s index (45); a measure which takes into account both species present and the relative abundance of species, and 4) Simpson evenness (46) to measure species representation. Phylogenetic beta-diversity measures such as weighted UniFrac (a quantitative measure that uses abundance to weight phylogenic tree branches) and unweighted UniFrac (a qualitative measure assessing unique phylogenetic branch fractions) (47), and nonphylogenetic Bray–Curtis dissimilarity (48) and Euclidean diversity (49) to assess between-sample differences in abundance per count were also conducted.

#### 2. Glucoregulatory outcomes

Fasting serum glucose concentrations were measured at BL and W6 using a YSI 2300 STAT Plus (Yellow Springs, Ohio). A 2-hour oral glucose tolerance test (OGTT) was conducted at BL and W6 using a 75g glucose drink (Fisherbrand™ Glucose Tolerance Test Beverage).

Whole blood glucose was measured prior to consumption of the OGTT drink (time 0) and 60 and 120 minutes after using an Accu-Chek Performa glucometer with capillary blood collected from a finger stick. Participants were instructed to finish the drink within five minutes of opening the bottle. OGTT area under the curve (AUC) over 120 minutes was calculated using the logarithmic trapezoidal method (50). Fasting insulin was assessed using enzyme-linked immunosorbent assay (Millipore Cat.# EZHI-14K). Quantitative insulin-sensitivity check index (QUICKI), an estimate of fasting insulin sensitivity, was computed using [1/[log(fasting insulin uU/ml) + log(fasting glucose mg/dL)] (51). Homeostatic model assessment for insulin resistance (HOMA-IR), an estimate of fasting insulin resistance was computed using [fasting insulin (µU/L) × fasting glucose (nmol/L)/22.5] (52). HOMA-beta, an estimate of insulin secretion, was computed using [360 × fasting insulin(µU/mL) / fasting glucose (mg/dL)-63] (53)

#### 3. Cardiovascular outcomes

Resting systolic and diastolic BP were measured at BL and W6 using an OMRON automatic BP device (model BP:725, Kyoto, Japan) on the left arm after allowing participants to sit quietly for five minutes. Fasting lipid profile (LDL, HDL, total cholesterol, and triglycerides) were measured at BL and W6 using an Alere Cholestech LDX (San Diego, CA) device with blood obtained from a fingerstick and collected with a capillary tube.

#### 4. Body composition outcomes

All anthropometric and body composition measurements were obtained at BL and W6.

Height was measured in centimeters via a stadiometer. Weight was measured via a standard scale. Segmental body composition was obtained using a bioelectrical impedance device (RJL Systems Quantum – V Segmental, Ref: Q5S). All participants were instructed to be well-hydrated to obtain accurate measurements. Waist circumference was measured at the narrowest part of the abdomen, Hip circumference was measured at the widest part of the hips-gluteal region.

#### 5. Diet and physical activity assessments

A Diet History Questionnaire (DHQIII) was completed to establish a clear understanding of dietary patterns and specific food consumption over the previous month at BL and W6 (54). A triaxial accelerometer (model: wGT3X-BT) was used to assess free living physical activity over three days (two weekday and one weekend period) at BL and W6.

### Statistical analysis

The differential abundance of the ASVs at the different taxonomic levels, namely, phylum, class, family, order, genus, and species and differential abundance of pathways was analyzed using the limma-voom pipeline in R (version 4.0.4) (55). Only taxa and pathways detected in at least 25% of the samples were included in downstream analyses. A group, week, and group × week interaction model was fit, and participant id was treated as a random effect using the duplicate Correlation function. The week × group effect was adjusted for multiple testing (false discovery rate) within the taxa levels and pathways using the Benjamini–Hochberg procedure (56). When statistically significant (Q-value<0.25) week × group effects were observed, contrasts between groups at different time points were extracted from the model and adjusted for multiple comparisons using the Benjamini–Hochberg procedure. For those variables demonstrating Q-value<0.25, baseline-adjusted analyses to determine adjusted group effects at week 6 were also condcuted. Effects of selected dietary variables were also tested on the taxa and pathway variables by including dietary variables as covariates in the models.

The beta-diversity distance/dissimilarity matrices were analyzed for the effects of week, group, and week × group using the permutational multivariate ANOVA (PERMANOVA) (57) package in Primer (version 7) (58).

For the alpha-diversity, anthropometric, body composition, glucoregulatory, cardiovascular, and dietary data, a linear mixed model analysis with nlme package (59) was conducted in R with week (BL and W6) as the within subject fixed effect, group (CT and RT) as the between subject fixed effect, the week × group interaction, and participant as the random effect. For the strength data, a linear mixed model analysis was conducted with week (BL and W6) as the within subject fixed effect and participant as the random effect. For the physical activity data, a mixed ANOVA analysis was conducted with rstatix package (60) in R with week (BL and W6) and day (weekday and weekend) as the within subject fixed effects, group (CT and RT) as the between subject fixed effect, and the week × group, group × day, and week × day × group interactions included. Data not meeting normality assumptions were transformed using the Johnson’s family of transformations (61), however, only the non-transformed data are presented for interpretation purposes unless otherwise indicated. Baseline-adjusted analyses to determine adjusted group effects at week 6 were also conducted. When statistically significant (p<0.05) week × group effects were observed, contrasts between groups at different time points were constructed and adjusted for multiple comparisons using the multivariate t distribution (mvt) adjustment in R (62).

For taxa demonstrating statistically significant differences over time by group, their associations with cardiometabolic outcomes were examined using Pearson’s correlation in JMP Pro (16.0.0) (63). The correlation coefficient were compared between groups using Fishers r to z transformation and statistical significance was assessed for the observed z test statistic.

The best set of cardiometabolic variables associated with the community structure (i.e., ASV abundance) was deduced using the BIOENV (64) analysis in the vegan package of R (65) as described previously (34). The statistical significance for the BIOENV procedure was assessed using the Mantel test (65). The input dataset for this procedure comprised: weight, BMI, FFM%, fat%, torso fat%, fasting glucose, glucose AUC_120 min_, fasting insulin, HOMA-IR, QUICKI, systolic BP, diastolic BP, waist circumference, total cholesterol, HDL, LDL, and triglycerides. The best subsets of variables for the RT and CT groups at BL and W6 were plotted as vectors along nonmetric, multidimensional scaling (NMDS) plots. The significance of the variables on the 2-dimensional ordinations was assessed using vegan function envfit (65). The envfit results describe the contribution of the variables to the ordinations and do not supersede the results of the BIOENV analyses.

## III. RESULTS

### Gut microbiome outcomes

#### Alpha diversity

Significant group × week interaction effects were observed for Shannon entropy, Simpson index, and Simpson evenness (group × week, p<0.05). Post-hoc pairwise analyses indicate that these effects were driven by significantly higher levels of Shannon entropy and Simpson index in RT compared to CT at BL, and significant increase in Simpson evenness index over 6 weeks in CT (mvt-adjusted pairwise p<0.05). Baseline adjusted analyses indicate higher Simpson index in CT compared with RT at W6 (BL adjusted group effect, p<0.05). The alpha-diversity results are presented in **Figure 2** and **Table S1**.

**Figure 2.**
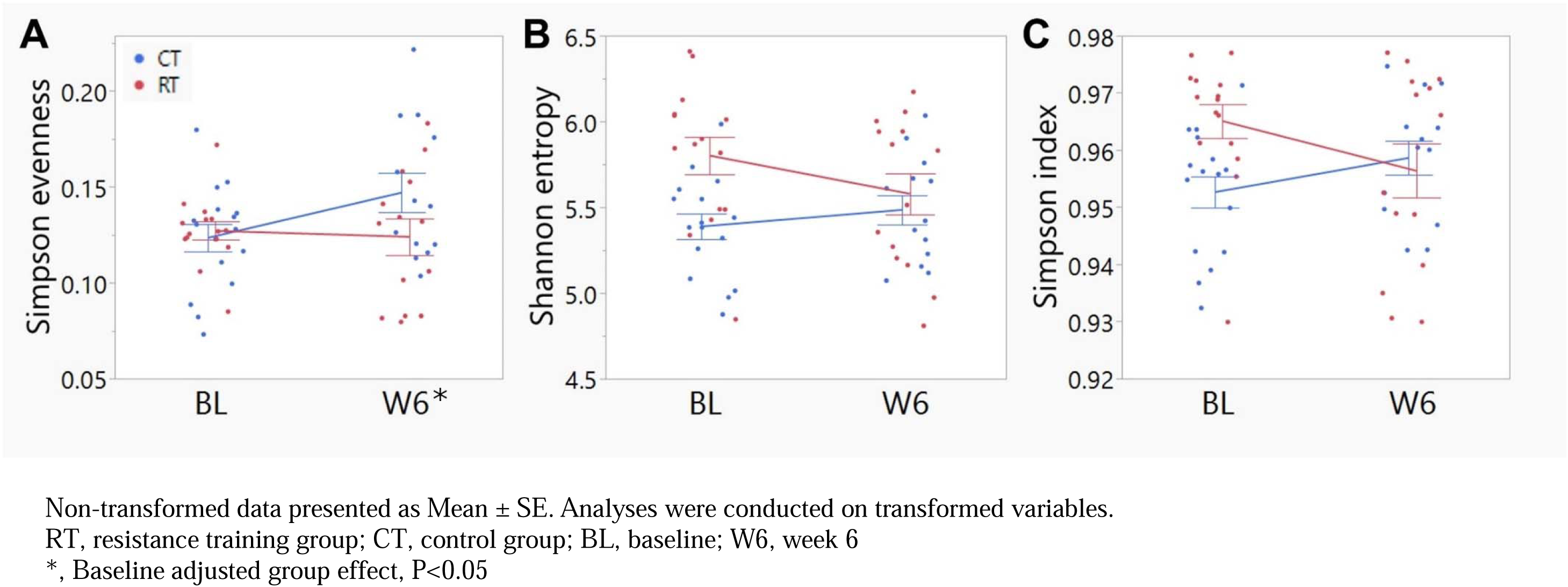
Indexes of alpha-diversity: (A) Simpson Evenness, (B) Shannon Entropy, and (C) Simpson Index between groups at BL and W6. Non-transformed data presented as Mean ± SE. Analyses were conducted on transformed variables. RT, resistance training group; CT, control group; BL, baseline; W6, week 6 *, Baseline adjusted group effect, P<0.05

#### Beta diversity

There was a significant group × week interaction effect only for Euclidian beta diversity (group × week effect, p<0.05), however post hoc results indicate no significant differences between group and week comparisons. Beta-diversity results for Euclidean, Bray-Curtis, Jaccard, weighted and unweighted unifrac matrices are presented in **Table S2**. The beta-diversity plots for Euclidean, weighted unifrac, and unweighted unifrac diversity are depicted in **Figure 3**.

**Figure 3.**
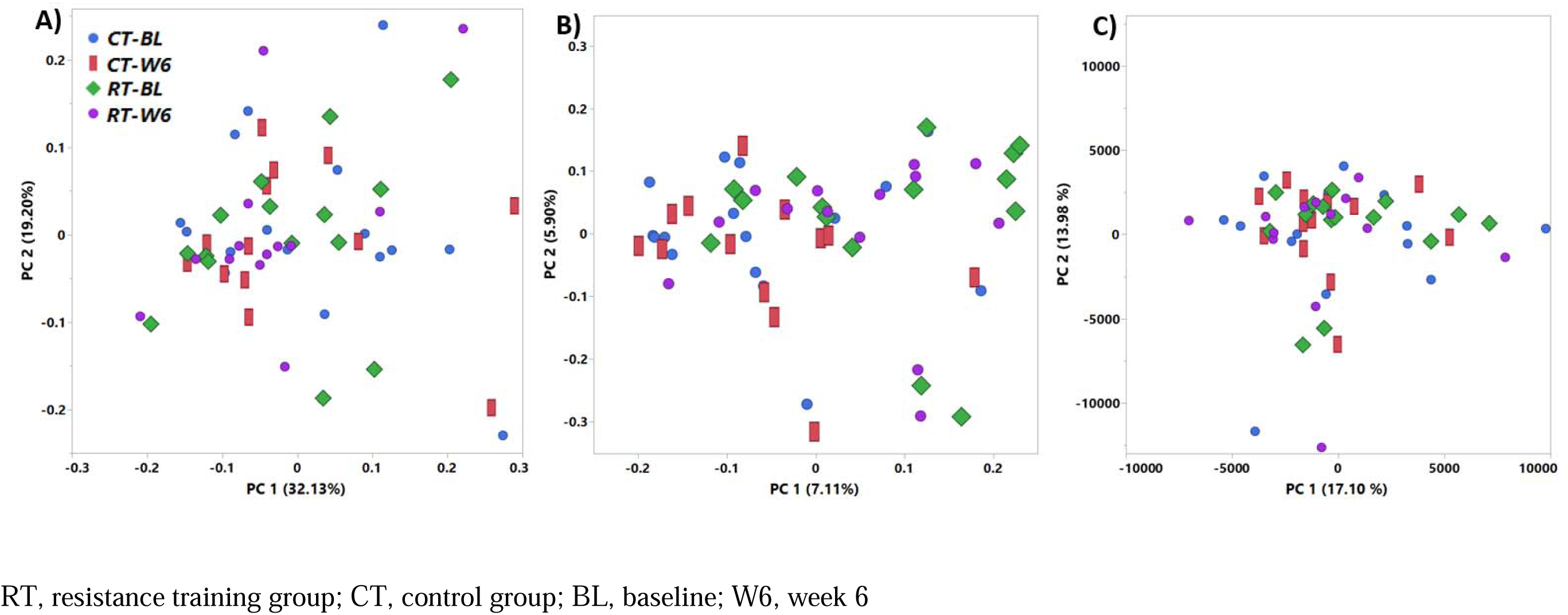
Beta-diversity measures (A) Weighted Unifrac, (B) Unweighted Unifrac, and (C) Euclidean between groups at BL and W6. RT, resistance training group; CT, control group; BL, baseline; W6, week 6

#### Taxa

Significant group × week interaction effects (p<0.05 and q<0.25) for the *Roseburia* genus, *Roseburia faecis*, and single ASV in *Faecalibacterium prausnitzii*, and in the *Roseburia* and *Holdemania* genera were observed. Post hoc pairwise analyses demonstrate significant increases in the *Roseburia* genus and *Roseburia faecis* over 6 weeks in RT, higher abundance of *Roseburia* genus ASV in RT compared to CT at week 6, increase in *Holdemania* ASV in CT over 6 weeks (BH-adjusted pairwise analysis, p<0.05), and a notable but non-significant increase in *Faecalibacterium prausnitzii* ASV over 6 weeks in RT (BH-adjusted pairwise analysis, p<0.1). Group × week interaction effects (p<0.05 but q> 0.25) were also detected for numerous other taxa and the relative abundances and results are presented in **Table 2**.

**Table 2.**
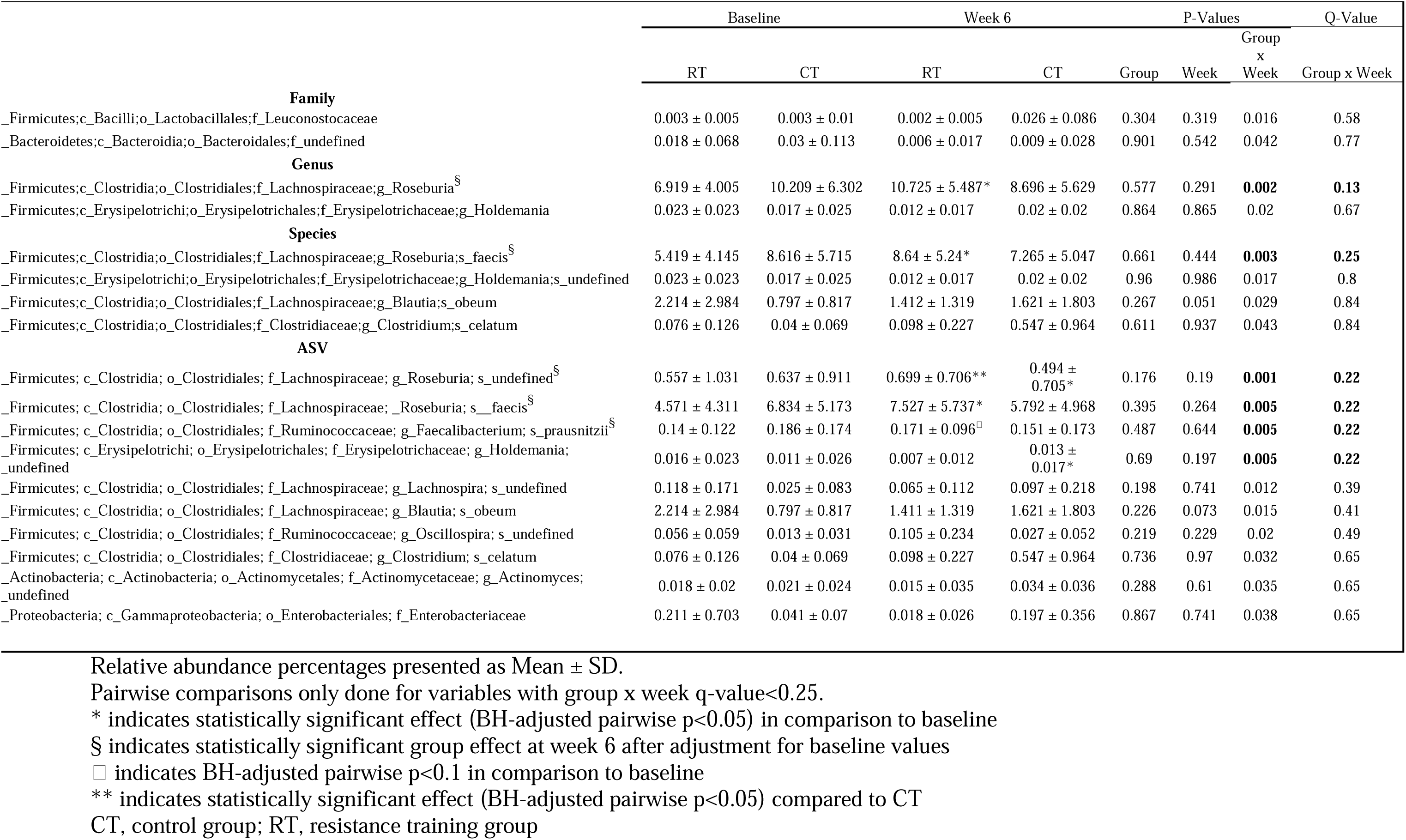
Microbial taxonomic relative abundance for RT and CT at baseline and week 6.

#### KEGG pathways

Numerous pathways demonstrated significant group × week interaction effects (p<0.05 and q<0.25), 1) carbohydrate metabolism i.e., starch and sucrose metabolism, ascorbate and aldrate metabolism, and pyruvate metabolism, 2) metabolism of other amino acids: phosphonate and phosphinate metabolism, 3) cellular processes i.e., flagellar assembly and bacterial chemotaxis, 4) genetic information processing: sulfur relay system, and 5) environmental adaptation: plant pathogen interaction. In the carbohydrate metabolism category, starch and sucrose metabolism was lower in RT compared to CT at BL and increased with RT over 6 weeks while ascorbate and aldrate metabolism was higher in CT at W6 compared to RT (BH-adjusted pairwise p<0.05). In cellular processes category, flagellar assembly and bacterial chemotaxis decreased with CT over 6 weeks and flagellar assembly was higher in RT compared to CT at W6 (BH-adjusted pairwise p<0.05). The genetic information processing category revealed that sulfur relay system decreased over 6 weeks in CT (BH-adjusted pairwise p<0.05). And the plant pathogen interaction pathway (environmental adaptation category) decreased with CT over 6 weeks (BH-adjusted pairwise p<0.05). Group × week interaction effects (p<0.05 but q>0.25) were also detected for numerous other pathways and the relative abundances and results are presented in **Table 3**.

**Table 3.**
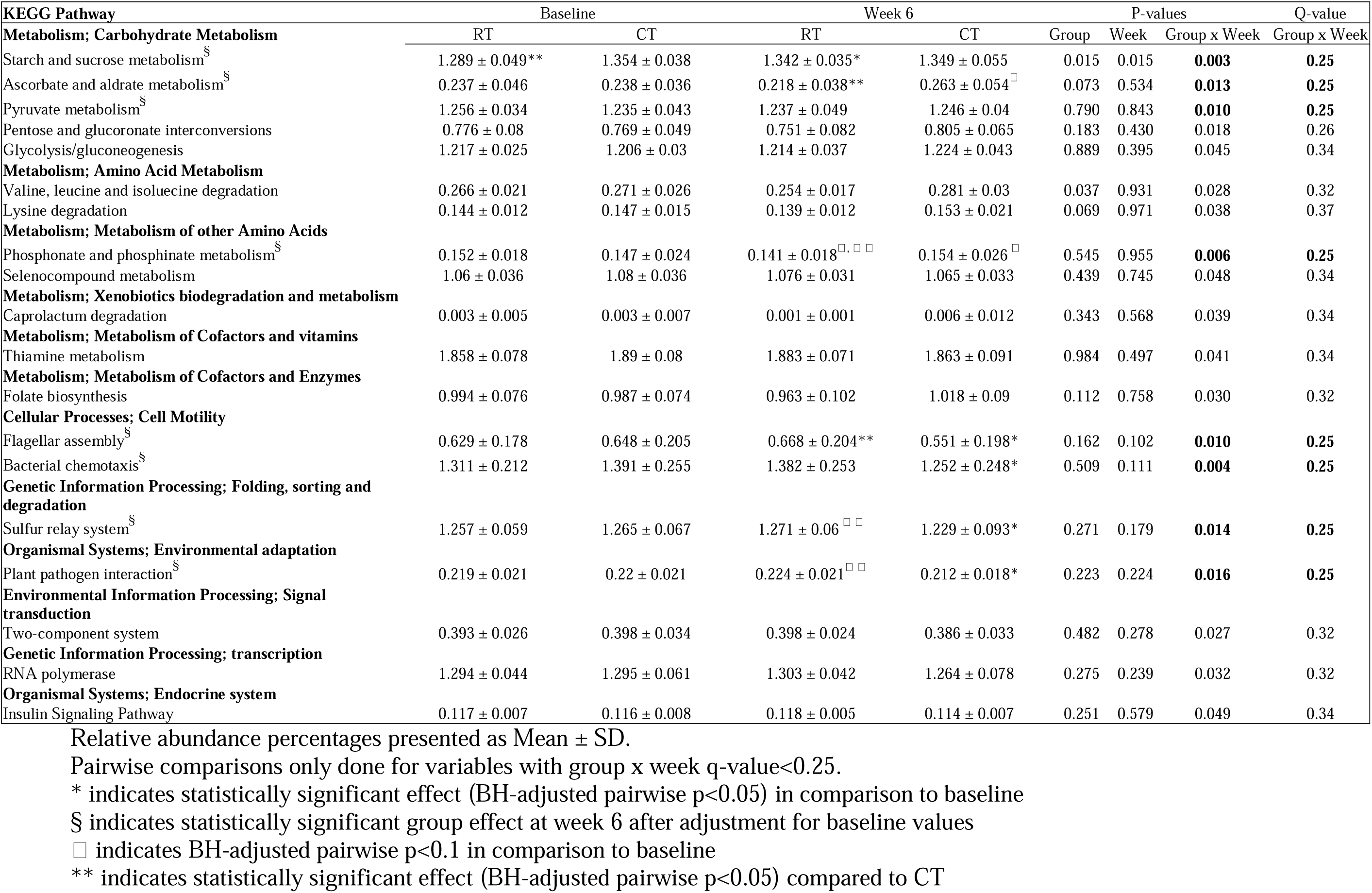
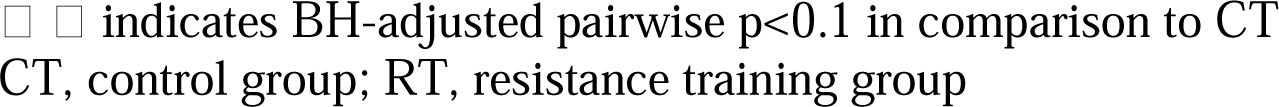
Microbial functions for RT and CT at baseline and week 6.

| KEGG Pathway | Baseline |  | Week 6 |  | P-values |  |  | Q-value |
| --- | --- | --- | --- | --- | --- | --- | --- | --- |
|  | RT | CT | RT | CT | Group | Week | Group x Week | Group x Week |
| **Metabolism; Carbohydrate Metabolism** |  |  |  |  |  |  |  |  |
| Starch and sucrose metabolism<sup>§</sup> | 1.289 ± 0.049** | 1.354 ± 0.038 | 1.342 ± 0.035* | 1.349 ± 0.055 | 0.015 | 0.015 | **0.003** | **0.25** |
| Ascorbate and aldrate metabolism<sup>§</sup> | 0.237 ± 0.046 | 0.238 ± 0.036 | 0.218 ± 0.038** | 0.263 ± 0.054<sup>□</sup> | 0.073 | 0.534 | **0.013** | **0.25** |
| Pyruvate metabolism<sup>§</sup> | 1.256 ± 0.034 | 1.235 ± 0.043 | 1.237 ± 0.049 | 1.246 ± 0.04 | 0.790 | 0.843 | **0.010** | **0.25** |
| Pentose and glucuronate interconversions | 0.776 ± 0.08 | 0.769 ± 0.049 | 0.751 ± 0.082 | 0.805 ± 0.065 | 0.183 | 0.430 | 0.018 | 0.26 |
| Glycolysis/gluconeogenesis | 1.217 ± 0.025 | 1.206 ± 0.03 | 1.214 ± 0.037 | 1.224 ± 0.043 | 0.889 | 0.395 | 0.045 | 0.34 |
| **Metabolism; Amino Acid Metabolism** |  |  |  |  |  |  |  |  |
| Valine, leucine and isoleucine degradation | 0.266 ± 0.021 | 0.271 ± 0.026 | 0.254 ± 0.017 | 0.281 ± 0.03 | 0.037 | 0.931 | 0.028 | 0.32 |
| Lysine degradation | 0.144 ± 0.012 | 0.147 ± 0.015 | 0.139 ± 0.012 | 0.153 ± 0.021 | 0.069 | 0.971 | 0.038 | 0.37 |
| **Metabolism; Metabolism of other Amino Acids** |  |  |  |  |  |  |  |  |
| Phosphonate and phosphinate metabolism<sup>§</sup> | 0.152 ± 0.018 | 0.147 ± 0.024 | 0.141 ± 0.018<sup>□, □ □</sup> | 0.154 ± 0.026<sup>□</sup> | 0.545 | 0.955 | **0.006** | **0.25** |
| Selenocompound metabolism | 1.06 ± 0.036 | 1.08 ± 0.036 | 1.076 ± 0.031 | 1.065 ± 0.033 | 0.439 | 0.745 | 0.048 | 0.34 |
| **Metabolism; Xenobiotics biodegradation and metabolism** |  |  |  |  |  |  |  |  |
| Caprolactum degradation | 0.003 ± 0.005 | 0.003 ± 0.007 | 0.001 ± 0.001 | 0.006 ± 0.012 | 0.343 | 0.568 | 0.039 | 0.34 |
| **Metabolism; Metabolism of Cofactors and vitamins** |  |  |  |  |  |  |  |  |
| Thiamine metabolism | 1.858 ± 0.078 | 1.89 ± 0.08 | 1.883 ± 0.071 | 1.863 ± 0.091 | 0.984 | 0.497 | 0.041 | 0.34 |
| **Metabolism; Metabolism of Cofactors and Enzymes** |  |  |  |  |  |  |  |  |
| Folate biosynthesis | 0.994 ± 0.076 | 0.987 ± 0.074 | 0.963 ± 0.102 | 1.018 ± 0.09 | 0.112 | 0.758 | 0.030 | 0.32 |
| **Cellular Processes; Cell Motility** |  |  |  |  |  |  |  |  |
| Flagellar assembly<sup>§</sup> | 0.629 ± 0.178 | 0.648 ± 0.205 | 0.668 ± 0.204** | 0.551 ± 0.198* | 0.162 | 0.102 | **0.010** | **0.25** |
| Bacterial chemotaxis<sup>§</sup> | 1.311 ± 0.212 | 1.391 ± 0.255 | 1.382 ± 0.253 | 1.252 ± 0.248* | 0.509 | 0.111 | **0.004** | **0.25** |
| **Genetic Information Processing; Folding, sorting and degradation** |  |  |  |  |  |  |  |  |
| Sulfur relay system<sup>§</sup> | 1.257 ± 0.059 | 1.265 ± 0.067 | 1.271 ± 0.06<sup>□ □</sup> | 1.229 ± 0.093* | 0.271 | 0.179 | **0.014** | **0.25** |
| **Organismal Systems; Environmental adaptation** |  |  |  |  |  |  |  |  |
| Plant pathogen interaction<sup>§</sup> | 0.219 ± 0.021 | 0.22 ± 0.021 | 0.224 ± 0.021<sup>□ □</sup> | 0.212 ± 0.018* | 0.223 | 0.224 | **0.016** | **0.25** |
| **Environmental Information Processing; Signal transduction** |  |  |  |  |  |  |  |  |
| Two-component system | 0.393 ± 0.026 | 0.398 ± 0.034 | 0.398 ± 0.024 | 0.386 ± 0.033 | 0.482 | 0.278 | 0.027 | 0.32 |
| **Genetic Information Processing; transcription** |  |  |  |  |  |  |  |  |
| RNA polymerase | 1.294 ± 0.044 | 1.295 ± 0.061 | 1.303 ± 0.042 | 1.264 ± 0.078 | 0.275 | 0.239 | 0.032 | 0.32 |
| **Organismal Systems; Endocrine system** |  |  |  |  |  |  |  |  |
| Insulin Signaling Pathway | 0.117 ± 0.007 | 0.116 ± 0.008 | 0.118 ± 0.005 | 0.114 ± 0.007 | 0.251 | 0.579 | 0.049 | 0.34 |

Relative abundance percentages presented as Mean ± SD.
Pairwise comparisons only done for variables with group x week q-value<0.25.
\* indicates statistically significant effect (BH-adjusted pairwise p<0.05) in comparison to baseline
§ indicates statistically significant group effect at week 6 after adjustment for baseline values
□ indicates BH-adjusted pairwise p<0.1 in comparison to baseline
\*\* indicates statistically significant effect (BH-adjusted pairwise p<0.05) compared to CT
□ □ indicates BH-adjusted pairwise $p < 0.1$ in comparison to CT CT, control group; RT, resistance training group

### Anthropometric and body composition outcomes

Waist circumference, FM%, FFM% and torso FM% demonstrated significant group × week effect interaction effects (p<0.05). Post hoc pairwise comparisons indicate that the effect observed in waist circumference was driven by a significant increase in CT at W6 compared to BL (mvt-adjusted pairwise p<0.05). The baseline adjusted analyses indicate that FM%, torso FM%, and waist circumference were significantly higher and FFM% lower at W6 in CT compared to RT (BL adjusted group effect, p<0.05). Results are shown in **Table 4**.

**Table 4.**
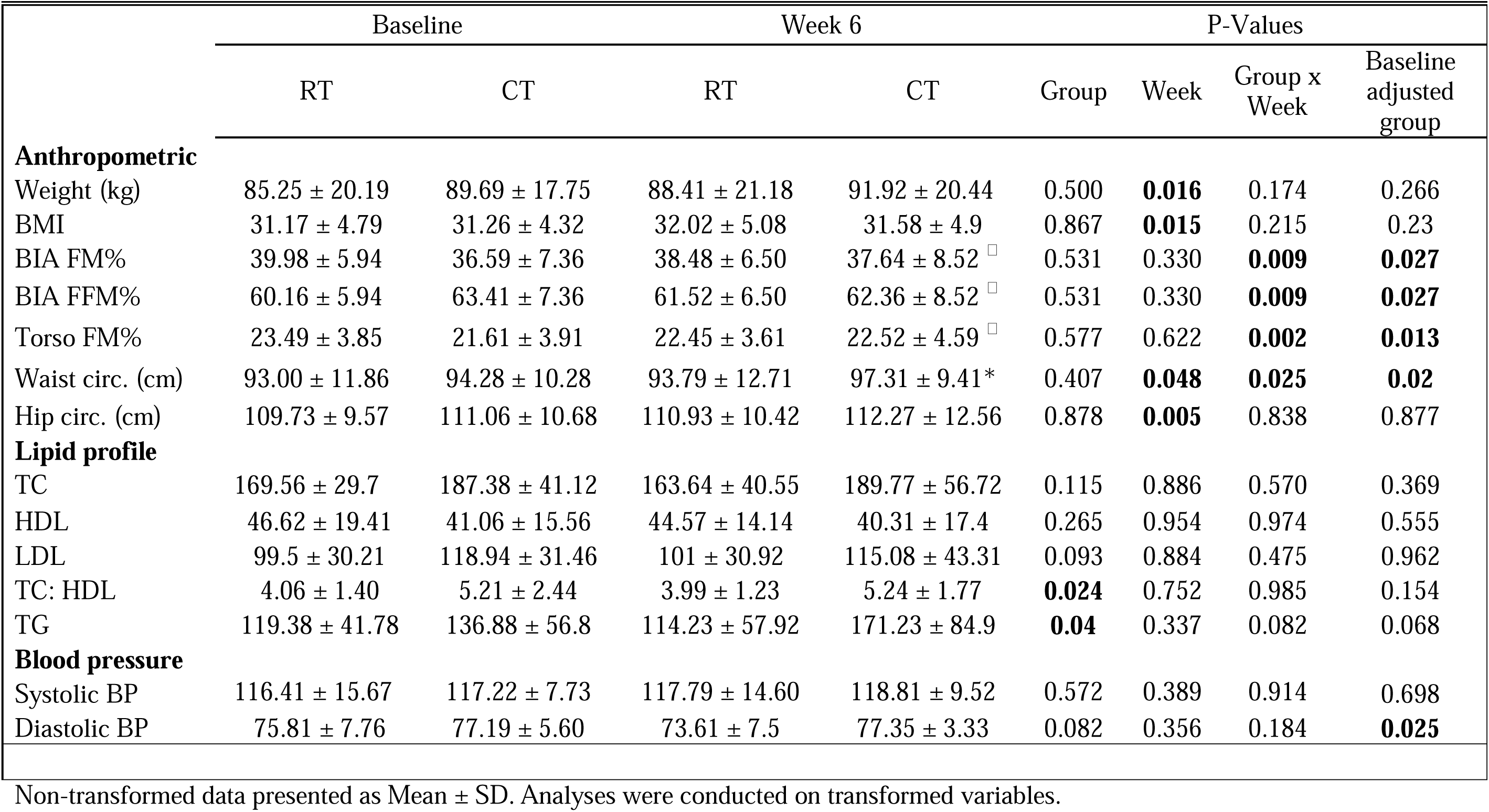
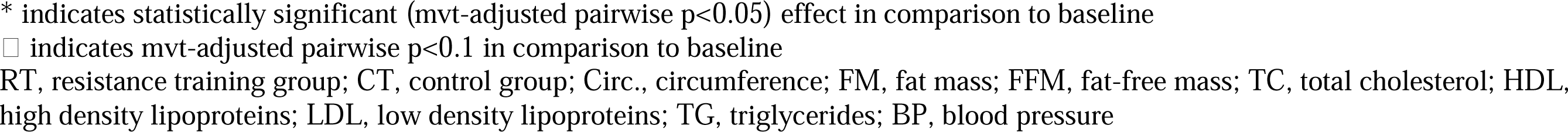
Anthropometric, body composition, and cardiovascular data for RT and CT at baseline and week 6.

|  | Baseline |  | Week 6 |  | P-Values |  |  |  |
| --- | --- | --- | --- | --- | --- | --- | --- | --- |
|  | RT | CT | RT | CT | Group | Week | Group x Week | Baseline adjusted group |
| **Anthropometric** |  |  |  |  |  |  |  |  |
| Weight (kg) | 85.25 ± 20.19 | 89.69 ± 17.75 | 88.41 ± 21.18 | 91.92 ± 20.44 | 0.500 | **0.016** | 0.174 | 0.266 |
| BMI | 31.17 ± 4.79 | 31.26 ± 4.32 | 32.02 ± 5.08 | 31.58 ± 4.9 | 0.867 | **0.015** | 0.215 | 0.23 |
| BIA FM% | 39.98 ± 5.94 | 36.59 ± 7.36 | 38.48 ± 6.50 | 37.64 ± 8.52 □ | 0.531 | 0.330 | **0.009** | **0.027** |
| BIA FFM% | 60.16 ± 5.94 | 63.41 ± 7.36 | 61.52 ± 6.50 | 62.36 ± 8.52 □ | 0.531 | 0.330 | **0.009** | **0.027** |
| Torso FM% | 23.49 ± 3.85 | 21.61 ± 3.91 | 22.45 ± 3.61 | 22.52 ± 4.59 □ | 0.577 | 0.622 | **0.002** | **0.013** |
| Waist circ. (cm) | 93.00 ± 11.86 | 94.28 ± 10.28 | 93.79 ± 12.71 | 97.31 ± 9.41* | 0.407 | **0.048** | **0.025** | **0.02** |
| Hip circ. (cm) | 109.73 ± 9.57 | 111.06 ± 10.68 | 110.93 ± 10.42 | 112.27 ± 12.56 | 0.878 | **0.005** | 0.838 | 0.877 |
| **Lipid profile** |  |  |  |  |  |  |  |  |
| TC | 169.56 ± 29.7 | 187.38 ± 41.12 | 163.64 ± 40.55 | 189.77 ± 56.72 | 0.115 | 0.886 | 0.570 | 0.369 |
| HDL | 46.62 ± 19.41 | 41.06 ± 15.56 | 44.57 ± 14.14 | 40.31 ± 17.4 | 0.265 | 0.954 | 0.974 | 0.555 |
| LDL | 99.5 ± 30.21 | 118.94 ± 31.46 | 101 ± 30.92 | 115.08 ± 43.31 | 0.093 | 0.884 | 0.475 | 0.962 |
| TC: HDL | 4.06 ± 1.40 | 5.21 ± 2.44 | 3.99 ± 1.23 | 5.24 ± 1.77 | **0.024** | 0.752 | 0.985 | 0.154 |
| TG | 119.38 ± 41.78 | 136.88 ± 56.8 | 114.23 ± 57.92 | 171.23 ± 84.9 | **0.04** | 0.337 | 0.082 | 0.068 |
| **Blood pressure** |  |  |  |  |  |  |  |  |
| Systolic BP | 116.41 ± 15.67 | 117.22 ± 7.73 | 117.79 ± 14.60 | 118.81 ± 9.52 | 0.572 | 0.389 | 0.914 | 0.698 |
| Diastolic BP | 75.81 ± 7.76 | 77.19 ± 5.60 | 73.61 ± 7.5 | 77.35 ± 3.33 | 0.082 | 0.356 | 0.184 | **0.025** |

Non-transformed data presented as Mean ± SD. Analyses were conducted on transformed variables.
\* indicates statistically significant (mvt-adjusted pairwise $p < 0.05$ ) effect in comparison to baseline
□ indicates mvt-adjusted pairwise $p < 0.1$ in comparison to baseline
RT, resistance training group; CT, control group; Circ., circumference; FM, fat mass; FFM, fat-free mass; TC, total cholesterol; HDL, high density lipoproteins; LDL, low density lipoproteins; TG, triglycerides; BP, blood pressure

### Cardiovascular outcomes

Baseline adjusted analyses indicated that diastolic BP was lower in RT compared to CT at W6 (BL adjusted group effect, p<0.05). Results are shown in **Table 4**.

### Glucoregulatory outcomes

QUICKI demonstrated significant group × week interaction effects (p<0.05). Post hoc pairwise analysis indicates a significant increase in QUICKI over 6 weeks in RT (mvt-adjusted pairwise p<0.05). Baseline adjusted analyses indicate higher fasting glucose in CT, higher QUICKI in RT, and lower HOMA-IR in RT at W6 (BL adjusted group effect, p<0.05). Results are shown in **Figure 4** and **Table S3.**

**Figure 4.**
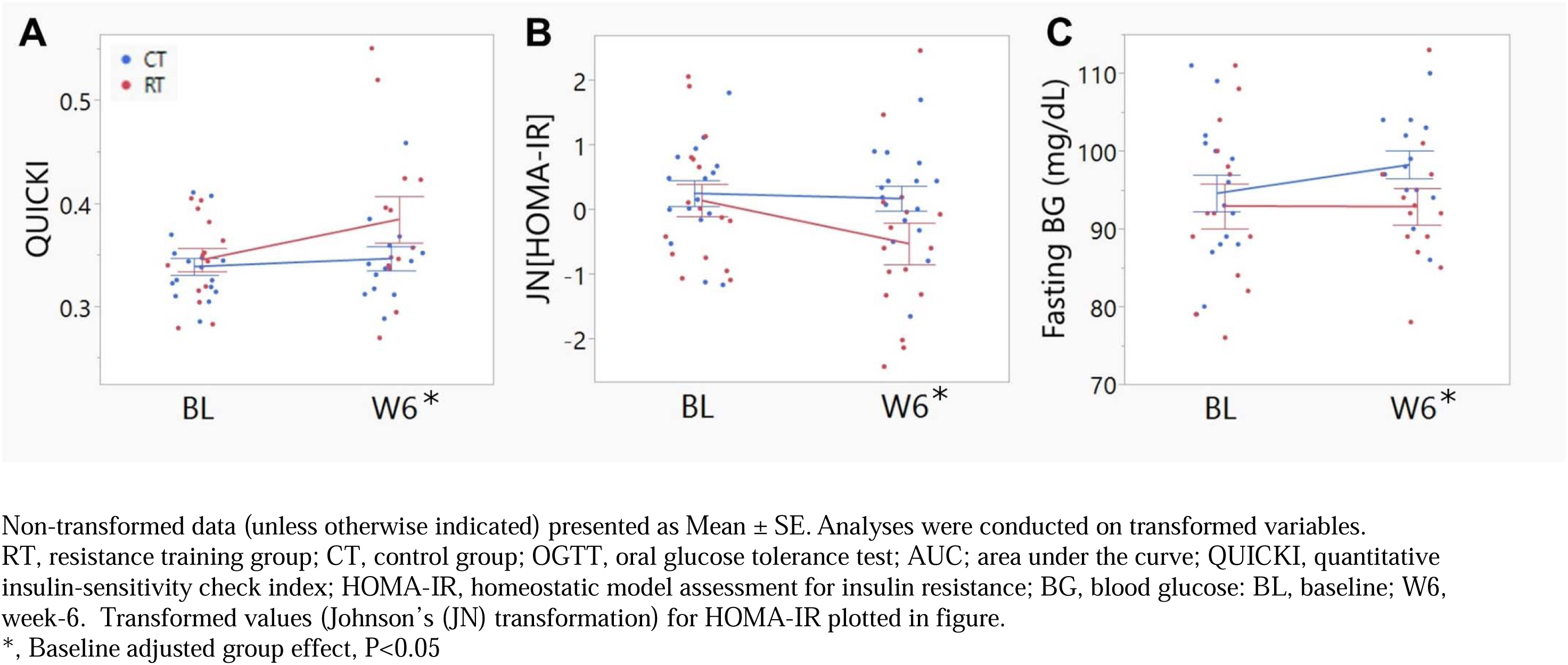
Glucoregulatory measures (A) QUICKI, (B) HOMA-IR, and (C) Fasting BG between groups at BL and W6. Non-transformed data (unless otherwise indicated) presented as Mean ± SE. Analyses were conducted on transformed variables. RT, resistance training group; CT, control group; OGTT, oral glucose tolerance test; AUC; area under the curve; QUICKI, quantitative insulin-sensitivity check index; HOMA-IR, homeostatic model assessment for insulin resistance; BG, blood glucose: BL, baseline; W6, week-6. Transformed values (Johnson’s (JN) transformation) for HOMA-IR plotted in figure. *, Baseline adjusted group effect, P<0.05

### Strength gains in RT

Strength significantly increased in bench max, row max, squat max, push up test, and plank test in the RT group over 6 weeks (week effect, p<0.05, **Table S4**). There were no strength assessments conducted for the CT group.

### Physical activity

Percentage of time spent in sedentary activity, percentage of time spent in light activity, and total time of sedentary bouts demonstrated significant 3-way group × week × day interaction effects (p<0.05). Post-hoc tests for the variables did not reveal any significant or meaningful differences. There were no significant interaction effects observed for total energy expenditure (kcals) or METs between groups over the course of the intervention. Data and results are presented in **Table S5.**

### Dietary intake

Pinitol demonstrated significant week × group interaction effects (p<0.05, **Table S6**). Pairwise analyses indicate pinitol was significantly higher in CT compared to RT at W6 (mvt-adjusted pairwise p<0.05). Baseline adjusted analyses indicate higher intake pinitol and sorbitol in CT compared to RT at W6 (BL adjusted group effect, p<0.05, **Table S6**). Moreover, inclusion of pinitol and sorbitol, in addition to dietary carbohydrates, total sugars, and dietary fiber as covariates, did not affect the significant week x group interaction effects of the aforementioned microbial taxa and pathway.

### Association of microbial community structure with cardiometabolic variables

The best correlated subset of cardiometabolic variables with the microbial community structure in the RT group at BL (model Spearman rho=0.43; p=0.009), CT group at BL (model Spearman rho=0.14; p=0.16), RT group at W6 (model Spearman rho=0.40; p=0.01), and CT group at W6 (model Spearman rho=0.35; p=0.027) are depicted in **Figure 5**. The subset of cardiometabolic variables with the highest correlation with the community structure comprised of FFM%, fat%, fasting glucose concentrations, and fasting insulin concentrations in RT at baseline (**Figure 5A**), fasting glucose in CT at BL which was not statistically significant (**Figure 5B**), diastolic blood pressure and BMI in RT at W6 (**Figure 5C**), systolic and diastolic BP, and LDL in CT at W6 (**Figure 5D**). Moreover, fasting glucose (R2=42%; p<0.05 envfit test), FFM% (R2=46%; p<0.05), and fat% (R2=46%; p<0.05) each demonstrated a statistically significant association with the 2-dimensional NMDS ordination of RT at BL.

**Figure 5.**
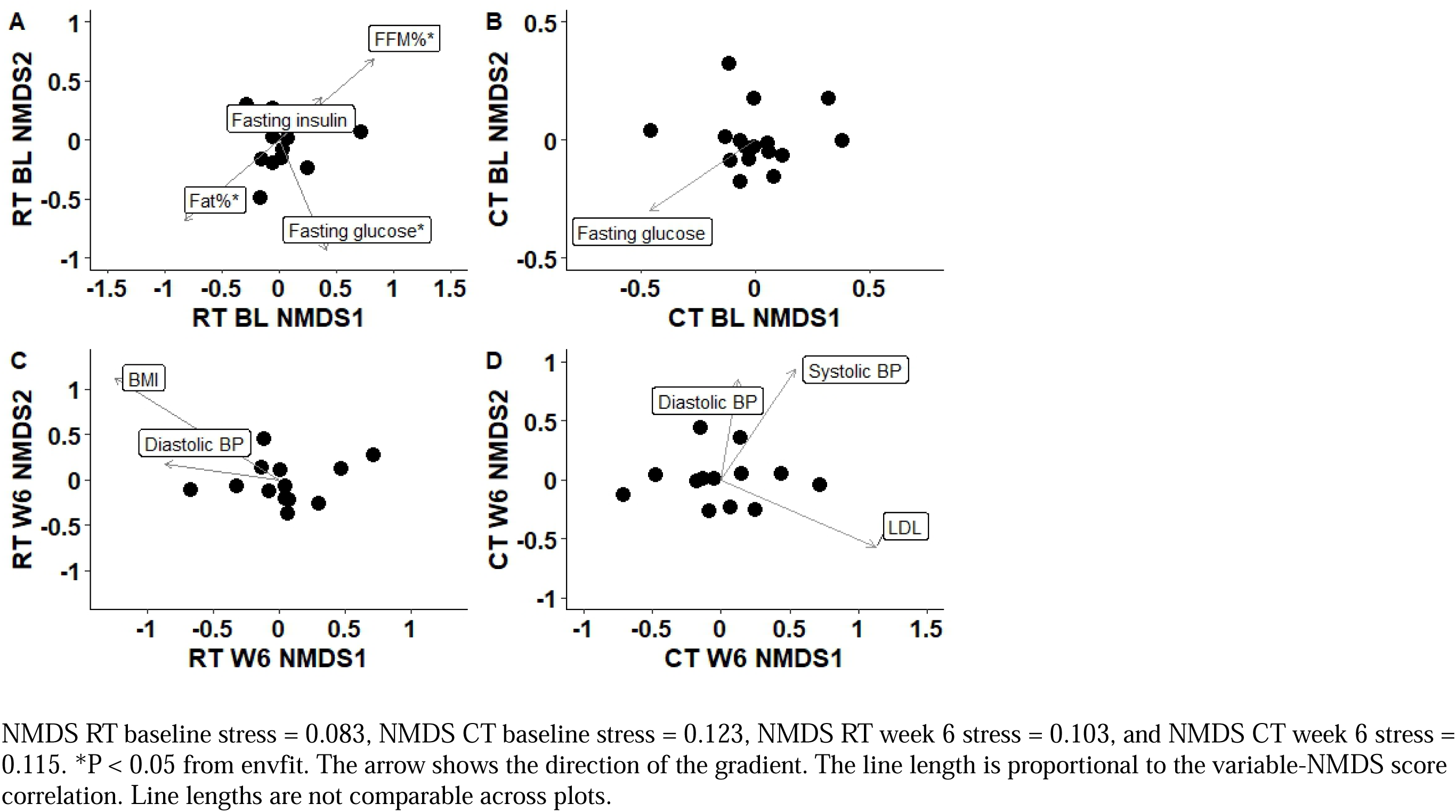
Nonmetric multidimensional scaling (NMDS) plot of the Bray–Curtis dissimilarities of microbial data with best set of clinical and dietary variables at (A) baseline in RT, (B) baseline in CT, (C) week 6 in RT, and (D) week 6 in CT. NMDS RT baseline stress = 0.083, NMDS CT baseline stress = 0.123, NMDS RT week 6 stress = 0.103, and NMDS CT week 6 stress = 0.115. *P < 0.05 from envfit. The arrow shows the direction of the gradient. The line length is proportional to the variable-NMDS score correlation. Line lengths are not comparable across plots.

### Correlation between Roseburia genus and glucoregulatory outcomes

Since the *Roseburia* genus was the only taxa to demonstrate a strong change with RT over 6 weeks in comparison to the CT group, we examined its association with cardiometabolic outcomes. At BL in the RT group, there was a modest positive correlation of *Roseburia* with diastolic BP (r=0.52, p<0.05, **Table S7)** which was not observed at W6. However, there were no statistically significant differences found for the *Roseburia*-Diastolic BP correlations between the RT and CT groups at either BL or W6.

There was a notable but non-significant moderately positive correlation between *Roseburia* and QUICKI (r=0.48, p<0.1, **Table S7**) in the RT group at W6. At BL, there were no statistically significant group differences found for the correlations of *Roseburia* with QUICKI, but the group differences were apparent at W6 (p<0.1 for RT vs. CT at W6, **Table S7**). There were no significant or notable correlations observed between *Roseburia* and body composition measures or glucose concentrations.

## IV. DISCUSSION

The RT intervention demonstrated notable effects on the gut microbiome and cardiometabolic outcomes. RT increased butyrate producers in the *Roseburia* genus and induced changes in several microbial pathways involved in carbohydrate metabolism and cell motility.

RT also had positive effects on insulin sensitivity indices which could potentially be associated with an increase in butyrate producers.

RT increased *Roseburia,* a genus attributed to butyrate production in the gut (66). A notable (albeit non-significant) increase in *Faecalibacterium prausnitzii*, another known producer of butyrate and overall indicator of a healthy gut (67), was also seen with RT. These findings are corroborated by cross-sectional studies, which have observed a higher prevalence of butyrate producers in individuals who regularly exercise compared to those leading sedentary lifestyles (68). Moreover, intervention studies conducted on individuals with diabetes and non-alcoholic fatty liver disease (NAFLD) have revealed an increased abundance of the Roseburia genus following aerobic exercise and combined aerobic and resistance exercise (69, 70).

However, there is very limited evidence from previous studies focusing solely on resistance training concerning the impact on Roseburia. Although SCFAs such as butyrate are commonly produced by microbial fermentation of non-digestible fibers (71), exercise increases gut motility and mucus secretion (68) which can alter the interaction among microbiota, thereby potentially impacting SCFA production. Enhanced butyrate production could thus be the result of butyrate producers such as *Roseburia* cross-feeding with mucin degraders (72), or the result of increased mucin colonization as several *Roseburia* species demonstrate mucin-adhering capability (73, 74). This increase in butyrate producers with RT has important clinical implications due to their roles in butyrate production and the observed effects of butyrate on the reduction of adiposity and insulin resistance (75).

Butyrate specifically has been shown to have protective effects against adiposity as well as insulin resistance, whether it be increased via direct supplementation or through endogenous gut production, the latter method being associated with other benefits such as increased gut endocrine function (75, 76). Our correlation analyses did not demonstrate any associations of adiposity with butyrate producers in the RT group most likely because the adiposity changes were largely driven by an increase in adiposity with CT over 6 weeks while RT demonstrated protective effects or marginal decreases in adiposity overall. The insulin sensitivity index was improved 8.6% following the RT intervention, agreeing with much of the previous literature demonstrating that exercise can reduce insulin resistance (77, 78). Sedentary behavior and obesity are both risk factors for the development of insulin resistance driven by impairments in multiple pathways related to insulin signaling. Exercise is known to attenuate these impairments via multiple mechanisms such as increased glucose clearance, increased skeletal muscle mass, increased insulin receptors, and increased GLUT4 translocation (79). These findings also support previous findings highlighting the protective effects of physical activity against the development of type 2 diabetes and metabolic syndrome (80). The increases in selected SCFA producers may be associated with improvements in insulin sensitivity. In fact, exercise responders i.e. individuals demonstrating greater than 2 times change in insulin sensitivity were found to have an increased capacity for SCFA production in men with prediabetes, (81) indicating the important association of SCFA with glucoregulation. In our study, this is supported by the correlation analyses that demonstrate a potential link between *Roseburia* and QUICKI suggesting that those with higher abundance of *Roseburia* also exhibited improved insulin sensitivity.

Although circulating SCFA were not measured in the present study, the effects of SCFA, specifically butyrate, on insulin sensitivity could be via multiple mechanisms. As reviewed previously, SCFA can activate G-protein coupled receptors on gut enteroendocrine L-cells which can stimulate the release of GLP-1, an incretin hormone, thereby promoting insulin secretion and decreasing glucagon secretion (82). SCFA may have additional effects on insulin sensitivity in skeletal muscle and the liver, two tissues responsible for glucose uptake. SCFA can lead to upregulation of AMPK activity, which stimulates mobilization of glucose transporters like GLUT4, leading to improvements in insulin sensitivity (82).

The microbial community structure was also found to be associated with blood pressure in the NMDS analyses. Specifically, RT resulted in lower diastolic blood pressure at the end of the 6-week intervention, which is indicative of an improvement of cardiovascular health. There is evidence to suggest that SCFAs can regulate BP by binding to G-protein coupled receptors GPR41, GPR43, and GPR109A which triggers a cascade of pathways inducing vasodilation and reducing BP (83). Although it is plausible to assume that the increases in butyrate producers with RT may serve as potential mechanisms for the observed improvements in diastolic blood pressure (84), our correlation analysis did not support these ideas.

Microbial pathways involved in carbohydrate metabolism such as starch and sucrose metabolism were differentially modulated with RT. The increased abundance of starch and sucrose metabolism pathways with RT appears to be independent of dietary changes as inclusion of carbohydrates, sugars, or dietary fiber as covariates did not change significance of these results. We speculate that the increased abundance of genes in these pathways with RT may be indicative of increased efficiency of microbial starch and sucrose metabolism at any given intake level i.e., the microbiome becomes more adept at metabolizing these carbohydrates by increasing the abundance of these genes. This is a plausible theory considering that the positive effects of exercise on host carbohydrate metabolism are well established (85, 86). Although previous studies have observed differences in microbial metabolic pathways, including carbohydrate and amino acid metabolism, the lack of a clear pattern may be attributed to variations in population characteristics such as prediabetes, type 2 diabetes, and exercise interventions (20, 81). To further investigate these findings, future studies employing meta-transcriptomics of fecal samples could confirm whether changes in metabolic pathway abundances translate to alterations in gene expression.

RT also induced higher microbial flagellar assembly, a pathway involved in cell motility, at the end of the 6-week intervention compared to CT. While there is no current association between exercise and cell motility, these increases may be indicative of a more active gut microbiome. One of the motility mechanisms of bacteria involve the use of flagella to navigate their environment (87). Flagellar assembly is a complex process that ultimately allows bacteria to swim through liquids or adhere to semi-solid surfaces (88). The higher flagellar assembly pathway abundance with RT may be indicative of an increased ability of bacteria to colonize the RT-induced gut environment.

The changes observed in alpha-diversity following RT are largely driven by the distinct differences observed between the RT and CT groups at baseline. Exercise studies that have examined alpha-diversity measures have presented mostly non-significant results indicating no effect of a true strength training intervention on diversity (27, 31). Studies comparing strength training to aerobic exercise training also report no changes in either alpha-or beta-diversity (27, 31), but these studies were also limited by the lack of control groups. The present study did not demonstrate appreciable changes in beta-diversity with RT. However, there have been reports of changes in beta-diversity following strength training and increased protein intake in humans, but the results of strength training alone cannot be isolated in that study (26) due to the lack of an RT only group. Moreover, changes in beta-diversity are not always positive changes and are often reported without reference to the direction of change. The subject of exercise and microbial diversity still warrants further attention.

Several mechanisms have been proposed for the effects of exercise on the gut microbiome (89). Exercise-induced heat stress and intestinal permeability may acutely result in direct contact between microbes and the immune system, thereby changing microbial architecture and activity (89). Exercise can also increase abdominal mechanical forces, reduce GI transit time, and alter gut motility which can impact mucus secretion, intestinal pH, and nutrient availability (89). Moreover, acute exercise can lead to reduction in mesenteric blood flow to the gut resulting in localized intermittent hypoxia (90, 91), which could acutely promote a more anaerobic gut environment potentially promoting increased inhabitation of anaerobic bacteria, such as species in the *Roseburia* genus (92), a genus that increased following RT in the present study. Additionally, we speculate that the metabolites and hormones that are increased in the blood immediately following exercise (i.e., lactate, cortisol, etc.) (93, 94) could potentially be transported to the gut once at-rest blood flow is returned. The exposure of these metabolites, hormones, and cytokines to the gut could provide additional stimuli for the gut microbiome and lead to an environmental shift that could promote and/or deter the inhabitation of various microbes in the gut. Hence, the beneficial impact of RT on gut microbial taxa and functions could be via long-term adaptations resulting from these mechanisms and should be explored in future studies.

This study has numerous strengths and provides significant findings that contribute to a larger body of literature in the field of exercise and the gut microbiome. To our knowledge, this is one of the first studies to look solely at the effects of RT without any additional dietary or lifestyle intervention in humans and with the inclusion of a rigorous control. This study highlighted not only alterations in the gut microbiome, but potential associations between SCFA producers and insulin sensitivity which should be examined further. In the context of real-world applications and viability for the studied intervention to be utilized as a treatment method, all participants who began the RT intervention (came to the first session) completed the entire intervention, speaking to the plausibility of this studied intervention to be adapted and safely utilized by this population outside of a research setting. Additionally, this intervention was specifically designed to reflect a program designed by a certified personal trainer to promote not just physiological and cardiometabolic changes, but also functional strength, mobility, and balance.

The limitations of this study pertain to not controlling for other lifestyle factors, hence some variability in the control group data was also observed, which is a known challenge in human clinical studies. Diet composition is known to have a significant effect on the gut microbiome as well as other markers of cardiometabolic health and adiposity (95). While diet was assessed before and after the intervention to ensure that no major shifts occurred over the course of the intervention, and participants were asked to consume diets similar to their baseline diets 3 days before their week 6 visit, no other method of controlling the diet was used, which could be a potential confounder on inter-individual microbiome variability. Evidence also indicates that other lifestyle factors such as sleep can have significant implications for adiposity and insulin resistance (96) and potentially on the gut microbiome (97) but was not assessed due to the scope of this study. Additionally, it is important to note that this study was conducted in individuals with overweight and obesity, so any observations in this study should not be applied to leaner populations as previous research has demonstrated differing effects of exercise between lean and obese populations (7). Other limitations are related to not collecting multiple stool samples each at baseline and week 6 to minimize temporal variability due to time of stool collection, and lack of direct SCFA assessment to provide important context to the data.

Additionally, we used predictive modeling for assessing microbial functions. However, PICRUST2 (42) is a common tool for predicting microbial functions based on 16S marker genes, and demonstrates strong correlations (0.79-0.88) for prediction of KO genes when compared to the gold-standard shotgun metagenomics sequencing (42).

This study provides preliminary evidence that resistance training increases abundance of selected SCFA producers and microbial metabolic pathways and improves cardiometabolic health. Future studies should 1) interrogate the microbiome-SCFA-glucoregulation axis in response to exercise with larger sample sizes, and 2) the interaction of lifestyle factors i.e., diet, exercise, and sleep on the gut microbiome in a controlled manner to gain a greater understanding of the complex relationship of the gut microbiome with human physiology and metabolism.

## Data Availability

All data produced in the present study are available upon reasonable request to the authors

