## Supplemental Table and Figures for "The Effects of 6 Weeks of Resistance Training on the Gut Microbiome and Cardiometabolic Health in Young Adults with Overweight and Obesity"

**Table S1.** Microbial alpha diversity for RT and CT at baseline and week 6.

|  | Baseline | | Week 6 | | P-Values | | | |
| --- | --- | --- | --- | --- | --- | --- | --- | --- |
|  | RT | CT | RT | CT | Group | Week | Group x Week | Baseline adj. group |
| Shannon entropy | 5.80 ± 0.42** | 5.39 ± 0.30 | 5.58 ± 0.45 | 5.49 ± 0.31 | **0.020** | 0.263 | **0.014** | 0.600 |
| Simpson index | 0.97 ± 0.01** | 0.95 ± 0.01 | 0.96 ± 0.02 | 0.96 ± 0.01 | 0.077 | 0.906 | **0.002** | 0.085 |
| Simpson evenness | 0.13 ± 0.02 | 0.12 ± 0.03 | 0.12 ± 0.04 | 0.15 ± 0.04* | 0.202 | 0.086 | **0.025** | **0.032** |
| CHAO 1 index | 243.60 ± 58.47 | 183.75 ± 38.41 | 211.36 ± 39.36 | 178.08 ± 34.73 | **<0.001** | **0.023** | 0.173 | 0.550 |

Non-transformed data presented as Mean ± SD. Analyses were conducted on transformed variables.

* indicates statistically significant (mvt-adjusted pairwise p<0.05) effect in comparison to BL
** indicates statistically significant effect (mvt-adjusted pairwise p<0.05) compared to CT

RT, resistance training group; CT, control group

**Table S2.** Microbial beta diversity effects for RT and CT over the 6-week study.

|  | Mean distance/dissimilarity between groups | | | | P-Value (Perm) | | | Psuedo-F |
| --- | --- | --- | --- | --- | --- | --- | --- | --- |
|  | RT: BL - W6 | CT: BL - W6 | BL: RT - CT | W6: RT - CT | Group | Week | Group x Week | Group x Week |
| Bray Curtis | 0.6 | 0.64 | 0.65 | 0.63 | 0.535 | 0.486 | 0.513 | 0.999 |
| Euclidean | 11546 | 11925 | 12035 | 11878 | 0.804 | 0.978 | **0.003** | 2.453 |
| Jaccard | 0.8 | 0.81 | 0.83 | 0.82 | 0.554 | 0.471 | 0.498 | 1 |
| Unweighted Unifrac | 0.66 | 0.65 | 0.68 | 0.66 | 0.545 | 0.481 | 0.51 | 0.999 |
| Weighted Unifrac | 0.27 | 0.28 | 0.28 | 0.28 | 0.532 | 0.502 | 0.48 | 1.001 |

RT, resistance training group; CT, control group; W6, week 6; BL, baseline

**Table S3.** Glucoregulatory data for RT and CT at baseline and week 6.

|  | Baseline | | Week 6 | | P-Values | | | |
| --- | --- | --- | --- | --- | --- | --- | --- | --- |
|  | RT | CT | RT | CT | Group | Week | Group x Week | Baseline adjusted group |
| Fasting glucose (mg/dL) | 92.93 ± 10.67 | 94.56 ± 9.25 | 92.85 ± 8.44 | 98.23 ± 6.46 | 0.125 | 0.229 | 0.141 | **0.015** |
| Fasting insulin (μU/mL) | 15.48 ± 13.66 | 14.67 ± 9.42 | 12.27 ± 15.39 | 12.89 ± 8.49 | 0.254 | **0.011** | 0.085 | 0.073 |
| OGTT AUC | 15285 ± 1953.7 | 16355.63 ± 2525.83 | 14757.69 ± 2144.16 | 16278.46 ± 2981.81 | 0.052 | 0.296 | 0.343 | 0.137 |
| QUICKI | 0.35 ± 0.04 | 0.34 ± 0.03 | 0.38 ± 0.08* | 0.35 ± 0.04 | 0.16 | **0.009** | **0.041** | **0.035** |
| JN[HOMA-IR]^1^ | 0.13 ± 1 | 0.24 ± 0.79 | -0.54 ± 1.28 | 0.16 ± 0.78 | 0.193 | **0.014** | 0.055 | **0.048** |
| HOMA-Beta | 469.56 ± 457.84 | 386.1 ± 311.32 | 431.3 ± 1105.55 | 292.81 ± 202 | 0.68 | **0.026** | 0.235 | 0.313 |

Non-transformed data (unless otherwise indicated) presented as Mean ± SD. Analyses were conducted on transformed variables.

* indicates statistically significant (mvt-adjusted pairwise p<0.05) effect in comparison to baseline
RT, resistance training group; CT, control group; OGTT, oral glucose tolerance test; AUC; area under the curve; QUICKI, quantitative insulin-sensitivity check index; HOMA-IR, homeostatic model assessment for insulin resistance; HOMA-beta, homeostatic model assessment for beta cell function

^1^Transformed values (Johnson’s (JN) transformation) presented for HOMA-IR

**Table S4.** RT strength data at baseline and week 6

|  | RT | |  |
| --- | --- | --- | --- |
|  | Baseline | Week 6 | Week effect P-value |
| Bench max (lbs) | 96.6 ± 42.7 | 112.9 ± 45.1 | **<0.001** |
| Row max (lbs) | 107.3 ± 38.4 | 133.4 ± 48.7 | **<0.001** |
| Squat max (lbs) | 163.5 ± 60.0 | 213.0 ± 67.1 | **<0.001** |
| Pushup test (reps) | 9.7 ± 5.7 | 22.3 ± 6.7 | **<0.001** |
| Plank hold test (min.) | 50.1 ± 30.0 | 79.0 ± 26.2 | **<0.001** |

Data presented as Mean ± SD. Analyses were conducted on non-transformed variables.

All data presented as Mean ± SD
RT, resistance training group

**Table S5.** Free-living physical activity data for weekdays and weekend days in RT and CT at baseline and week 6.

|  | Baseline | | | | Week 6 | | | | P-Values | | |
| --- | --- | --- | --- | --- | --- | --- | --- | --- | --- | --- | --- |
|  | RT | | CT | | RT | | CT | | Week | Group x Day | Group x Week x Day |
|  | Weekday | Weekend | Weekday | Weekend | Weekday | Weekend | Weekday | Weekend |  |  |  |
| Energy expenditure (kcals) | 414.63 ± 121.34 | 494.06 ± 337.7 | 460.9 ± 235.83 | 506.91 ± 268.8 | 673.43 ± 711.92 | 1513.8 ± 2650.65 | 630.57 ± 419.94 | 431.2 ± 174.16 | **0.05** | 0.511 | 0.194 |
| METs | 1.22 ± 0.06 | 1.27 ± 0.18 | 1.24 ± 0.14 | 1.23 ± 0.14 | 1.27 ± 0.14 | 1.29 ± 0.18 | 1.34 ± 0.25 | 1.18 ± 0.09 | 0.185 | 0.153 | 0.201 |
| Step counts | 5796.54 ± 1436.71 | 5578.91 ± 3496.99 | 5601.67 ± 2665.32 | 5589.64 ± 2454.03 | 7151.61 ± 3429.24 | 6653.33 ± 2905.92 | 7369.5 ± 3284.67 | 5502 ± 1894.55 | **0.039** | 0.88 | 0.224 |
| Total time of sedentary bouts (min) | 236.6 ± 111.44 | 247.89 ± 165.21 | 288.61 ± 205.53 | 168.67 ± 88.25 | 262.27 ± 158.34 | 225.08 ± 140.76 | 243.34 ± 158.71 | 259.81 ± 147.32 | 0.533 | 0.552 | **0.004** |
| Sedentary activity (% time) | 79 ± 6 | 76 ± 12 | 79 ± 9 | 75 ± 8 | 77 ± 11 | 76 ± 11 | 76 ± 10 | 79 ± 8 | 0.209 | 0.971 | **0.046** |
| Light activity (% time) | 16 ± 5 | 19 ± 9 | 16 ± 7 | 20 ± 7 | 17 ± 11 | 18 ± 8 | 17 ± 7 | 17 ± 8 | 0.226 | 0.515 | **0.032** |
| Moderate activity (% time) | 4.9 ± 1.6 | 5.6 ± 3.9 | 4.8 ± 2.8 | 4.6 ± 2.5 | 6.2 ± 3.5 | 5.5 ± 3.3 | 6.3 ± 3.4 | 3.7 ± 2.1 | 0.285 | 0.235 | 0.202 |
| Vigorous activity (% time) | 0.1 ± 0.2 | 0.2 ± 0.2 | 0.2 ± 0.3 | 0.1 ± 0.1 | 0.2 ± 0.2 | 0.4 ± 0.5 | 0.6 ± 1.1 | 0.1 ± 0.2 | 0.197 | **0.023** | 0.094 |

Non-transformed data presented as Mean ± SD. Analyses were conducted on transformed variables.

All data presented as Mean ± SD.
CT, control group; RT, resistance training group

**Table S6.** Dietary intake for RT and CT at baseline and week 6.

|  | Baseline | | Week 6 | | P-Values | | | |
| --- | --- | --- | --- | --- | --- | --- | --- | --- |
|  | RT | CT | RT | CT | Group | Week | Group x Week | Baseline adjusted group |
| Total energy (kcal) | 1685.5 ± 739.9 | 1569.2 ± 647.1 | 1493.4 ± 556.3 | 1376.9 ± 554.1 | 0.469 | **0.007** | 0.97 | 0.615 |
| Carbohydrates (g) | 196.1 ± 94.1 | 184.5 ± 64.7 | 167.5 ± 62.5 | 158.3 ± 62.9 | 0.657 | **<0.001** | 0.964 | 0.794 |
| Carbohydrates (% of kcal) | 46.4 ± 8.5 | 49.8 ± 9.6 | 45.1 ± 7.4 | 46.5 ± 5.9 | 0.283 | 0.142 | 0.711 | 0.685 |
| Protein (g) | 70.8 ± 27.4 | 64.3 ± 33.2 | 62.6 ± 23.6 | 61.7 ± 33.2 | 0.488 | 0.132 | 0.555 | 0.921 |
| Protein (% of kcal) | 17.3 ± 3.2 | 16.2 ± 3.6 | 17.1 ± 3.5 | 17.1 ± 3.9 | 0.456 | 0.617 | 0.41 | 0.726 |
| Fat (g) | 69.2 ± 35.2 | 62.9 ± 33.5 | 63.1 ± 30.9 | 54.5 ± 20.8 | 0.234 | 0.142 | 0.461 | 0.68 |
| Fat (% of kcal) | 36.5 ± 5 | 34 ± 8.1 | 37 ± 5.1 | 36.5 ± 7.1 | 0.562 | 0.199 | 0.385 | 0.698 |
| Pinitol (g) | 0.006 ± 0.008 | 0.008 ± 0.01 | 0.002 ± 0.004** | 0.008 ± 0.007 | 0.057 | 0.989 | **0.034** | **0.004** |
| Sorbitol (g) | 0.311 ± 0.261 | 0.239 ± 0.162 | 0.22 ± 0.191 | 0.246 ± 0.127 | 0.889 | 0.899 | **0.05** | **0.041** |
| Starch (g) | 71.9 ± 37.4 | 75.2 ± 37.6 | 65.9 ± 28.5 | 61.5 ± 24.2 | 0.858 | 0.011 | 0.329 | 0.234 |
| Total sugars (g) | 93.3 ± 54.6 | 76 ± 35.5 | 75.2 ± 36.7 | 71.1 ± 48.8 | 0.491 | **0.027** | 0.217 | 0.405 |
| Total dietary fiber (g) | 15.97 ± 6.05 | 15.01 ± 6.84 | 13.38 ± 4.85 | 13.1 ± 5.79 | 0.543 | **0.006** | 0.479 | 0.815 |

Non-transformed data presented as Mean ± SD. Analyses were conducted on transformed variables.

All data presented as Mean ± SD.
** indicates statistically significant (mvt-adjusted pairwise p-value<0.05) effect compared to CT

CT, control group; RT, resistance training group

**Table S7.** Correlations of *Roseburia* genus with Diastolic BP and QUICKI at BL and W6 for RT and CT

| Group | Week | Variables | Pearson's correlation | Correlation P-Value | BL (CT vs. RT)  Z-Value | BL (CT vs. RT)  P-value |
| --- | --- | --- | --- | --- | --- | --- |
| CT | BL | Log10[*Roseburia*] - Log10[Diastolic BP] | 0.08 | 0.767 | -1.25 | 0.211 |
| RT | BL | Log10[*Roseburia*] - Log10[Diastolic BP] | 0.523 | 0.045 |  |  |
| CT | BL | Log10[*Roseburia*] - QUICKI | -0.118 | 0.662 | -0.31 | 0.757 |
| RT | BL | Log10[*Roseburia*] - QUICKI | 0.003 | 0.991 |  |  |
|  |  |  |  |  | W6 (CT vs. RT) Z-Value | W6 (CT vs. RT) P-Value |
| CT | W6 | Log10[*Roseburia*] - Log10[Diastolic BP] | -0.041 | 0.894 | -0.59 | 0.555 |
| RT | W6 | Log10[*Roseburia*] - Log10[Diastolic BP] | 0.214 | 0.462 |  |  |
| CT | W6 | Log10[*Roseburia*] - QUICKI | -0.279 | 0.356 | -1.92 | 0.055 |
| RT | W6 | Log10[*Roseburia*] - QUICKI | 0.503 | 0.067 |  |  |

BP, blood pressure; QUICKI, quantitative insulin-sensitivity check index

*
